# Age-related dysmodulation of systemic immune-inflammatory indices is associated with aggressive colorectal cancer in West Africa

**DOI:** 10.1101/2023.08.05.23293698

**Authors:** Jude Ogechukwu Okoye, Michael Emeka Chiemeka, Felix Emeka Menkiti, Eric Chukwudi Iheakwoaba, Nneka Agbakoba

## Abstract

**Introduction:** Despite the high mortality rate among colorectal cancer (CRC) patients in Africa, patients still bear the huge cost-related burden of cancer management. To reduce this burden, there is a current search for affordable markers for disease assessment and treatment monitoring. Contributing to this effort, this study evaluated systemic immune-inflammatory indices (SIII) among CRC patients.

**Methods:** This study included 89 patients with CRC diagnosed from Jan. 2016 to Dec. 2022. The patients were sub-grouped based on age and chemotherapy response. The neutrophil-to-lymphocyte ratio (NLR), lymphocyte-to-monocyte ratio (LMR), platelet-to-lymphocyte ratio (PLR), platelets-neutrophils-to-lymphocytes ratio (PNLR), and neutrophils-to-lymphocytes platelets ratio (NLPR) were assessed and analyzed accordingly. Significance was set at p< 0.05.

**Results:** The median age of the patients was 58.0 years. Metastatic and stage III/IV CRCs were prevalent among patients older than 50 years compared with patients aged 50 years or less. Among patients aged > 50 years, the pre-treatment (pre-T) to post-treatment (post-T) total white blood cell count (TWBC), neutrophils, monocytes, and NLPR significantly increased whereas the post-T lymphocyte count and LMR significantly declined (p< 0.05). Post-T TWBC count and LMR of patients aged > 50 years were 1.5 times higher and 2.4 times lower, respectively compared with the post-T values of patients who were 50 years old or less (p< 0.05). The post-T PNLR/NLPR and LMR were 2.7/2.3 times higher and 4 times lower among chemotherapy-naïve patients compared with the post-T values of chemotherapy-experienced patients, respectively (p< 0.05). The post-T NLR, PLR, and PNLR among chemoresistance. patients were 2.4, 2.3, and 1.5 higher than the post-T values of chemosensitive patients at p= 0.027, 0.015, and 0.022, respectively.

**Conclusion:** This study revealed a higher frequency of CRC and mortality risk among patients older than 50 years. It suggests that SIII could be used as a prognostic tool for CRC.

## Introduction

According to the GLOBACAN 2020 estimates, colorectal cancer (CRC) is the third most common cancer and the second leading cause of cancer-related deaths in the world.^1^ The average age-standardized incident rate per 100,000 (ASIR) of colon cancer (CC) in Europe, America, Asia, and Africa were 22.4, 13.0, 10.3, and 5.1, respectively.^1^ Comparison of incidence-to-mortality revealed lower case fatalities in countries with a high/very high human development index (HDI) compared with countries with a low/medium HDI; 43.6% vs 62.3%.^1,2^ Reasons for the difference between the two HDIs may be due to differences in the stage at presentation, access to healthcare facilities, and affordability of chemotherapy.^3^ Another reason of note is that CRCs in Africa are very aggressive and unusually metastatic.^4,5^ In Nigeria, for instance, evidence shows that 34%, 70%, and 96%, of CRCs are poorly differentiated, right-sided (RCC), and invasive, respectively.^3,6^ The RCCs are larger in size, more advanced, and poorly differentiated compared with the left CCs, and patients with RCCs are older.^7-9^ RCC patients have poorer overall survival (OS) and disease-free survival (DFS) rates compared with LCC patients.^9^ These factors reveal the importance of identifying affordable procedures and biomarkers for CRC that are alternatives to the expensive repeated imaging for high-mortality risk patients who are in low-resource settings. One such approach is precision medicine, in which the assessment of systemic immune-inflammatory (SIII) biomarkers has emerged as a promising option.^10^ High pretreatment inflammatory indices have been associated with both a greater risk of cancer relapse in radically resected tumours and shorter survival for cancer patients with metastatic disease.^11^

Systemic inflammation plays a vital role in promoting disease progression in many cancers. For instance, inflammatory cells increase vascular permeability, cancer cell adhesion, infiltration, stromal invasion, and metastasis.^11^ Circulating neutrophils and lymphocytes are pro- and anti-inflammatory, respectively. Neutrophils secrete cytokines and chemokines, which then stimulate the differentiation of megakaryocytes into platelets and promote cancer progression while lymphocytes promote cytotoxic immune response to cancer. Thus, circulating high neutrophil and low lymphocyte counts, especially post-chemotherapy, are independently associated with poor OS and progression-free survival (PFS), especially in metastatic CRC patients.^12^ The differentiation of monocytes into tumour-associated macrophages (TAMs) in the tumour microenvironment, evidenced by elevated monocyte count (> 300/mm^3^), has been attributed to poor OS, DFS, and cancer-specific survival (CSS) through tumour infiltration and metastasis. A low lymphocyte–monocyte ratio (LMR < 2.82), can reflect an active inflammation status and has been associated with high-grade tumours and worse OS and DFS and is more likely to be left-sided.^12^ Since age and geographical location of patients, and availability of resources are determiners of host survival, irrespective of disease stage at diagnosis,^11^ this study assessed the clinical utility and prognostic value of inflammation-related markers for better stratification of CRC patients in Southern Nigeria.

## Methods

### Study Population and Ethics

From January 2016 to December 2022, 98 patients with gastrointestinal diseases presented at the Department of Gastroenterology, Nnamdi Azikiwe University Teaching Hospital (NAUTH), Nigeria. Patients with inadequate records, especially hematological parameters (*n*□=□9) were excluded from the study. Finally, this study included a total of 89 patients diagnosed with CRC who were living in Anambra State. In addition to some antibiotics and surgeries, some patients received capecitabine and oxaliplatin as platinum chemotherapy. This retrospective study was approved by the NAUTH ethics committee (NAUTH/CS/66/VOL.15/VER.3/ 107/2022/081). The medical records of the patients were accessed for socio-clinical demographics such as age, gender, comorbidities, time of presentation, time of death, and contact for follow-up. All analyses were performed by the ethical standards laid down in the Declaration of Helsinki.

### Sample collection and handling

Peripheral whole blood samples (5 ml each) were collected into EDTA containers one week prior to the first chemotherapy and a week before discharge. Full blood counts were carried out on the whole blood samples using a Haemo-autoanalyzer. Following ultrasound investigations, biopsies or resected tissues were sent to the Department of Morbid Anatomy and Forensic Medicine for histological investigation. Two experienced pathologists evaluated the tissues for evidence of malignancy and metastasis. The neutrophils, lymphocytes, platelets, and monocyte counts were extracted along with their percentage of the total white cell count (10^^9^/L). Overall survival was calculated from the date of presentation or diagnosis to the date of death or last follow-up. The blood neutrophil-to-lymphocyte ratio (NLR), lymphocyte-to-monocyte ratio (LMR), platelet-to-lymphocyte ratio (PLR), platelets-neutrophils to lymphocytes ratio (PNLR; [Platelet count x Neutrophil count]/Lymphocyte count), and neutrophils-to-lymphocytes-platelets ratio (NLPR; [Neutrophil count x 100]/Lymphocyte count x platelet count]).

### Study design

The cases of CRC were categorized based on the following: 1. Age (≤ 50 years or > 50 years), 2. Condition on discharge (stable, unstable, and dead), 3. Time of symptom manifestation to the presentation at the clinic (TSMP; ≤ 6 months, 7 -12 months, and > 12 months), 4. platinum chemotherapy naïve or experienced (1-3 cycles and 4-6 cycles), 5. History of an herbal therapy (naïve or experienced), 6. Presence of metastasis, 7. Chemo-sensitive and chemo-resistance. Chemoresistance was determined by the assessment of features such as the alleviation of symptoms, liver and renal function tests, tumour response or degree of tumour shrinkage, and the need for second-line chemotherapy.^13^

### Statistical analysis

Chi-square/Fisher was used to determine the association between the socio-clinical demographics of patients who were ≤ 50 years and those > 50 years of age. Pearson’s correlation was used to determine the relationship between the variables (NLR, PLR, PNLR, NLPR, and LMR) before and after the last treatment. A T-test was used to compare data of 1. patients aged ≤ 50 years and > 50 years, 2. chemotherapy naïve and experienced patients, 3. patients who received 1-3 cycles and 4-6 cycles of chemotherapy, 4. herbal remedy experience and naïve patients, and 5. patients with and without metastatic tumours. ANOVA was used to compare data of patients who presented at ≤ 6 months, 7 -12 months, and > 12 months, and patients who were stable, unstable, and dead at discharge (in-hospital death). The overall survival of patients was analyzed using the Kaplan–Meier method. The survival probabilities between the subgroups were compared using the log-rank test.

## Result

The mean age, median age, and age range of the participants were 56.40 ± 13.58 years, 58.0 years, and 25 to 92 years, respectively. There was a high number of CRC diagnoses in 2018 (21.3%) compared with other years while a high rate of mortality was observed in 2020 (25.0%) and 2022 (25.0%) compared with other years (figure 1). The mean ‘time of symptom manifestation to presentation’ (TSMP) at the clinic was 11.8 ± 2.0 months (figure 2). The prevalent features presented by the patients were: Weight loss, constipation, intermittent diarrhea, low abdominal pain, and anorexia (Table 1a).

**Table 1a:** Features presented by CRC patients in Southern Nigeria.

| Variables | N= 89 | Percentage | Variables | N= 89 | Percentage |
| --- | --- | --- | --- | --- | --- |
| Weight loss | 59 | 67.0 | Rectal mass | 10 | 11.4 |
| Constipation | 57 | 64.8 | Dysuria | 10 | 11.4 |
| Diarrhea | 47 | 53.4 | Cough | 9 | 10.2 |
| Low Abdominal pain | 46 | 52.3 | Facal incontinence | 9 | 10.2 |
| Anorexia | 46 | 52.3 | Ascites | 9 | 10.2 |
| Hematochezia | 41 | 46.1 | Leg swelling | 9 | 10.2 |
| Pellet like stool | 38 | 43.2 | Abdominal mass | 8 | 9.2 |
| Tenesmus | 34 | 38.6 | Urinary retention | 8 | 9.1 |
| Abdominal distention | 33 | 37.5 | Fever | 8 | 9.1 |
| Fatigue | 23 | 26.4 | Nausea | 7 | 8.0 |
| Mucous stool | 23 | 26.1 | Nocturia | 6 | 6.8 |
| Vomiting | 23 | 26.1 | Hematuria | 6 | 6.8 |
|  |  |  | Post micturition |  |  |
| Melena | 16 | 18.2 | dribbling | 5 | 5.7 |
| Anal/Rectal bleeding | 15 | 17.0 | Fainting | 4 | 4.5 |
| Rectal prolapse | 15 | 17.0 | Jaundice | 3 | 3.4 |
| Dizziness | 12 | 13.6 | LUTs | 1 | 1.1 |
| Anal pain | 11 | 12.5 | Anemia | 1 | 1.1 |

**Figure 1:**
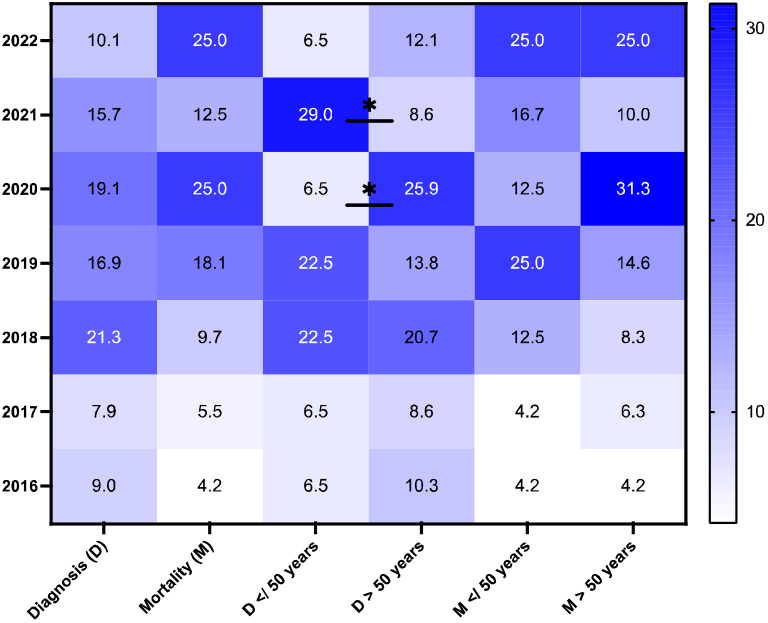
Heatmap of the percentage diagnosis and mortality rate among CRC patients age ≤ 50 years and > 50 years. Figure 1 shows that in 2020, the CRC diagnosis and mortality rates were approximately 4 times and 2.5 times higher among patients aged > 50 years compared with patients whose ages were less than or equal to (</; ≤) 50 years at p= 0.045 and 0.147, respectively. In 2021, the rate of CRC diagnosis was 3.4 times higher among patients who were ≤ 50 years old compared with those aged > 50 years at p= 0.016.

**Figure 2:**
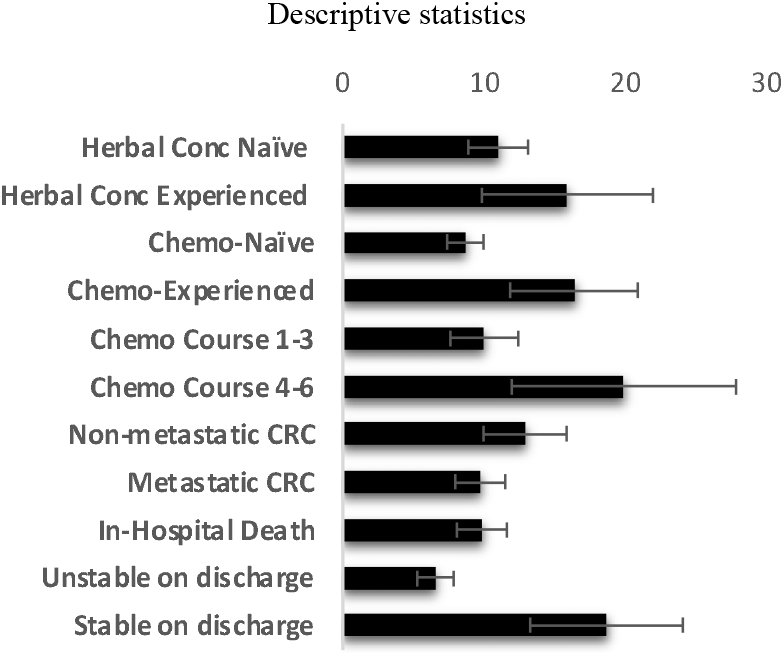
Time of symptom manifestation to presentation (Months) Figure 2 shows that herbal and chemotherapy naïve patients presented earlier than their experienced counterparts. Patients who received three or fewer courses of chemotherapy presented earlier than those who received more than four courses of chemotherapy. Patients with metastatic CRC presented at the clinic earlier than their counterparts with non-metastatic CRC. It also shows that patients who died in the hospital or unstable on discharge presented earlier than those who were stable on discharge.

### Age-related differences among CRC patients

The prevalence of CRC was slightly higher among females compared with men, especially among those who were under 50 years of age (table 1b). RCCs and metastasis were prevalent among patients who were over 50 years old whereas rectal tumours were prevalent in patients who were ≤ 50 years (p> 0.05). Less than 50% of the patients received chemotherapy and a higher percentage of those patients were under 50 years (p= 0.05). No significant difference was observed between the two age groups in terms of the history of herbal therapy use. The level of tertiary education was higher among patients who were ≤ 50 years compared with their over 50 years counterparts (p< 0.05). The history of tobacco use, alcohol consumption, and hypertension was prevalent among patients who were aged ≤ 50 years compared with their > 50 years counterparts (p> 0.05). Patients who were aged > 50 years old were approximately 1.8 times more like to present at the clinic in ≤ 6 months of symptom development compared with their ≤ 50 years counterparts (p= 0.046). The rate of unemployment was lower among patients who were older than 50 years compared with their ≤ 50 years counterparts (p> 0.05). The patients who were 50 years old and under had higher tumours grade than patients who were older than 50 years (p< 0.05). Based on histology, the prevalence of adenocarcinoma was higher among patients aged > 50 years compared with patients aged ≤ 50 years (p< 0.05). The rate of surgical resection uptake was higher among patients aged > 50 years (58.6%) compared with their ≤ 50 counterparts (54.8%) at p= 0.823. Post-T TWBC count was 1.5 times higher among patients who were above 50 years compared with their ≤ 50 years counterparts (p< 0.05) while the post-T monocyte and LMR were also lower among the former than the latter at p< 0.05 (table 2).

**Table 1b:** Socio-clinical characteristics of CRC patients in NAUTH.

| Variables | No. (%)<br>N= 89 | ≤ 50 years<br>n= 31 (%) | >50 years<br>n= 58 (%) | p- value |
| --- | --- | --- | --- | --- |
| **Sex** |  |  |  | 0.266 |
| Male | 43 (48.3) | 12 (38.7) | 31 (53.4) |  |
| Female | 46 (51.7) | 19 (61.3) | 27 (46.6) |  |
| **Employment status:** |  |  |  | 0.325 |
| Civil servant | 22 (24.7) | 7 (22.6) | 15 (25.9) |  |
| Dependant | 15 (14.6) | 3 (9.7) | 12 (20.7) |  |
| Self employed | 52 (58.4) | 21 (67.7) | 31 (53.4) |  |
| **Level of Education:** |  |  |  | 0.034* |
| No formal education | 5 (5.6) | 0 (0.0) | 5 (8.6) |  |
| Basic education | 52 (58.4) | 15 (45.5) | 37 (63.8) |  |
| Tertiary education | 32 (36.0) | 16 (50.0) | 16 (27.6) |  |
| **Alcohol consumption:** |  |  |  | 0.658 |
| No | 44 (49.4) | 14 (45.2) | 30 (51.7) |  |
| Yes | 45 (51.6) | 17 (54.8) | 28 (48.3) |  |
| **Tobacco Use:** |  |  |  | 0.746 |
| No | 77 (86.5) | 26 (83.9) | 51 (87.9) |  |
| Yes | 12 (13.5) | 5 (16.1) | 7 (12.1) |  |
| **History of Hypertension:** |  |  |  | 0.500 |
| No | 56 (62.9) | 18 (58.1) | 38 (65.5) |  |
| Yes | 33 (37.1) | 13 (41.9) | 20 (34.5) |  |
| **History of Herbal therapy:** |  |  |  | 0.999 |
| No | 77 (86.5) | 27 (87.1) | 50 (86.2) |  |
| Yes | 12 (13.5) | 4 (12.9) | 8 (13.8) |  |
| **TSMP:** |  |  |  | 0.077 |
| > 12 months | 17 (19.1) | 9 (29.1) | 8 (13.8) |  |
| 7-12 months | 33 (37.1) | 13 (41.9) | 20 (34.5) |  |
| ≤ 6 months | 39 (43.8) | 9 (29.0) | 30 (51.7) |  |
| **Tumour site:** |  |  |  | 0.698 |
| Right-sided Colon | 25 (28.1) | 7 (22.6) | 18 (31.0) |  |
| Left-sided Colon | 27 (30.3) | 10 (32.3) | 17 (29.3) |  |
| Rectum/Recto-sigmoid | 37 (41.6) | 14 (45.1) | 23 (39.7) |  |
| **Metastasis** |  |  |  | 0.002* |
| No | 59 (66.3) | 27 (87.1) | 32 (55.2) |  |
| Yes | 30 (33.7) | 4 (12.9) | 26 (44.8) |  |
| **Chemotherapy experience:** |  |  |  | 0.026* |
| No | 52 (58.4) | 13 (41.9) | 39 (67.2) |  |
| Yes | 37(41.6) | 18 (58.1) | 19 (32.8) |  |
| **Tumour grade** |  |  |  | 0.052* |
| Well differentiated | 37 (41.6) | 9 (29.0) | 28 (48.3) |  |
| Moderately differentiated | 43 (48.3) | 16 (51.6) | 27 (46.5) |  |
| Poorly differentiated | 9 (10.1) | 6 (19.4) | 3 (5.2) |  |
| **Histologic type** |  |  |  | 0.023* |
| Adenocarcinoma | 66 (74.2) | 19 (61.3) | 47 (81.0) |  |
| Squamous cell carcinoma | 17 (19.1) | 7 (22.6) | 10 (17.2) |  |
| Others | 6 (6.7) | 5 (16.1) | 1 (1.8) |  |
| **Disease Stage** |  |  |  | 0.006* |
| Stage 1 | 34 | 15 (48.4) | 19 (32.8) |  |
| Stage 2 | 25 | 12 (38.7) | 13 (22.4) |  |
| Stage 3 | 13 | 4 (12.9) | 9 (15.5) |  |
| Stage 4 | 17 | 0 (0.0) | 17 (29.3) |  |

TSMP: Time of symptom manifestation to presentation. Descriptive analysis and Chi-square/Fisher's exact test. \*Significance was set at $p < 0.05$ .

**Table 2:** Comparative analysis of hematological indices between two age groups.

| Variable | $\leq 50$ years<br>n= 31 | Fold (%)<br>change | $> 50$ years<br>n= 58 | Fold (%)<br>change | P -<br>value |
| --- | --- | --- | --- | --- | --- |
| | Mean ± SD | = AT/BT | Mean ± SD | = AT/BT | |
| WBC ($10^9/L$) BT | 8.38 ± 0.90 | | 8.37 ± 1.09 | | 0.580 |
| WBC ($10^9/L$) AT | 9.19 ± 1.50 | 1.10 (9.7) | 14.20 ± 4.50 | 1.70 (69.7) | 0.033* |
| P- value | 0.560 |  | 0.017* |  |  |
| Lymphocyte (%) BT | 36.40 ± 3.60 | | 34.12 ± 3.60 | | 0.203 |
| Lymphocyte (%) AT | 31.03 ± 4.11 | 0.85 (-14.8) | 27.05 ± 7.40 | 0.79 (-20.7) | 0.056 |
| P- value | 0.240 |  | 0.031* |  |  |
| Neutrophil (%) BT | 52.38 ± 4.09 | | 51.70 ± 5.05 | | 0.783 |
| Neutrophil (%) AT | 46.14 ± 7.54 | 0.88 (-11.9) | 57.33 ± 10.26 | 1.11 (10.9) | 0.138 |
| P- value | 0.287 |  | 0.016* |  |  |
| Platelet ($10^9/L$) BT | 322.03 ± 32.11 | | 405.08 ± 34.57 | | 0.979 |
| Platelet ($10^9/L$) AT | 376.42 ± 46.84 | 1.17 (16.9) | 330.44 ± 33.62 | 0.82 (-18.4) | 0.316 |
| P- value | 0.820 |  | 0.546 |  |  |
| Monocytes (%) BT | 7.23 ± 0.77 | | 3.33 ± 0.60 | | 0.138 |
| Monocytes (%) AT | 11.81 ± 3.57 | 1.63 (63.3) | 9.12 ± 2.33 | 2.74 (173.9) | 0.026* |
| P- value | 0.026* |  | 0.018* |  |  |
| NLR BT | 2.12 ± 0.36 | | 2.50 ± 0.56 | | 0.260 |
| NLR AT | 2.83 ± 0.74 | 1.33 (33.5) | 3.23 ± 1.07 | 1.29 (29.2) | 0.831 |
| P- value | 0.197 |  | 0.236 |  |  |
| PLR BT | 173.5 ± 23.83 | | 204.4 ± 37.98 | | 0.621 |
| PLR AT | 193.0 ± 55.00 | 1.11 (11.2) | 219.5 ± 45.51 | 1.07 (7.39) | 0.849 |
| P- value | 0.754 |  | 0.802 |  |  |
| PNLR BT | 830.63 ± 212.89 | | 1225.08 ± 232.1 | | 0.316 |
| PNLR AT | 953.90 ± 325.32 | 1.15 (14.8) | 1385.67 ± 211.3 | 1.13 (13.1) | 0.102 |
| P- value | 0.901 |  | 0.518 |  |  |
| NLPR BT | 0.84 ± 0.15 | | 0.57 ± 0.12 | | 0.307 |
| NLPR AT | 1.12 ± 0.32 | 1.33 (33.3) | 1.08 ± 0.33 | 1.89 (47.2) | 0.736 |
| P- value | 0.154 |  | 0.007* |  |  |
| LMR BT | 15.93 ± 4.01 | | 8.73 ± 2.23 | | 0.397 |
| LMR AT | 6.76 ± 3.92 | 0.42 (-57.6) | 2.87 ± 0.60 | 0.33 (-67.1) | 0.001* |
| P-value | 0.388 |  | < 0.001* |  |  |

Keys: BT; before treatment, AT; after treatment. Statistics: T-test. \*Significance was set at $p < 0.05$ .

Among patients aged ≤ 50 years, only the post-T monocyte significantly increased compared with the pre-T values at p< 0.05 (table 2). Among patients aged > 50 years, the post-T TWBC, neutrophils, monocytes, and NLPR values significantly increased whereas the post-T lymphocyte and LMR significantly reduced compared with the pre-T values (p< 0.05).

### Survival analysis

The overall mortality rate was 91.8%. Based on follow-up, the two-year and four-year survival rates of the patients were 5.2% and 3.9%, respectively. Chemo-experienced patients and patients who had non-metastatic tumours lived longer than chemo-naïve and patients with metastatic tumours (figure 3a). A significant inverse relationship was observed between metastasis and survival (p= 0.001). The in-hospital mortality rate was higher among cases with metastasis (46.2%) compared with non-metastatic cases (13.7%). Based on the survival rate, no significant difference was observed between chemo-experienced and herbal-experienced patients at p= 0.263. In Figure 3b, patients without a history of tobacco use, and alcohol consumption lived longer (162.7 days and 159.5 days) compared with the history (mean= 98.2 days and 135.8 days, respectively; p> 0.05). Patients who were both herbal and chemo-experienced had a higher mean survival rate (n= 2; 613 ± 76.50 days) compared with herbal/chemo-naïve patients (n= 42; 51.91 ± 14.61 days) at p< 0.001.

**Figure 3a:**
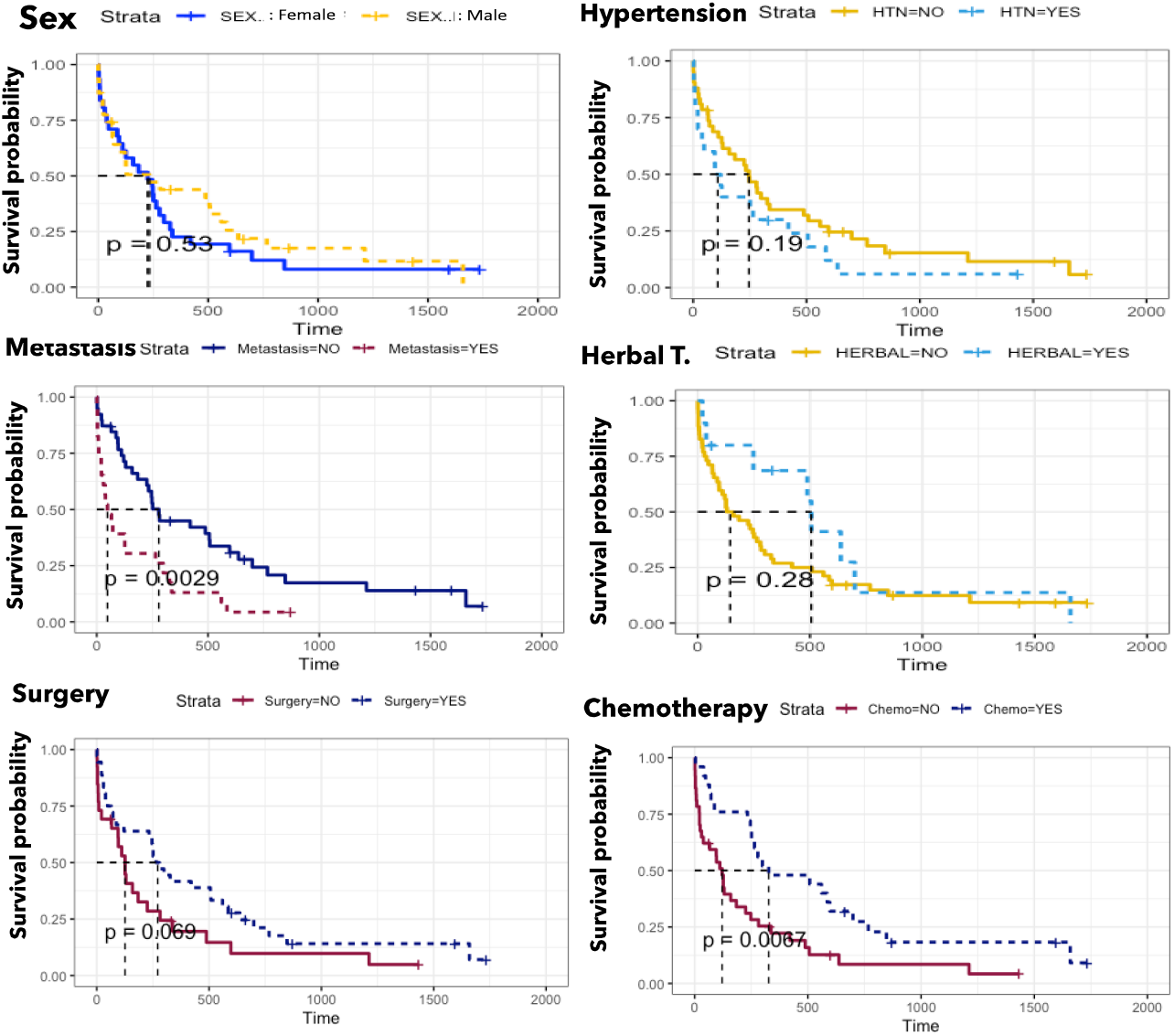
Survival analysis of CRC patients based on sex, history of hypertension and herbal therapy, uptake of surgery, chemotherapy, and occurrence of metastasis (Kaplan–Meier curve) Figure 3a shows that patients with non-metastatic CRC lived longer than their counterparts (mean= 594.6 days vs 268.2 days, respectively; p< 0.05). Those who were chemo-experienced also lived longer than their chemo-naïve counterparts (mean= 524.3 days vs 205.2 days, respectively; p< 0.05). Patients who had surgical resections lived longer than their surgery naïve counterparts (mean= 487.8 days vs 280.7 days, respectively; p> 0.05). More so, normotensive patients lived longer compared with patients with a history of hypertension (mean= 454.1 days vs 288.0 days, respectively; p> 0.05). The figure also shows that males lived longer than their female counterparts (mean/median= 462.2/233 days vs 344.8/225 days, respectively; p> 0.05). Interestingly, patients with a history of herbal therapy lived longer compared with herbal therapy-naïve patients (mean/median= 581.3/506.0 days vs 374.2/145 days, respectively; p> 0.05).

**Figure 3b:**
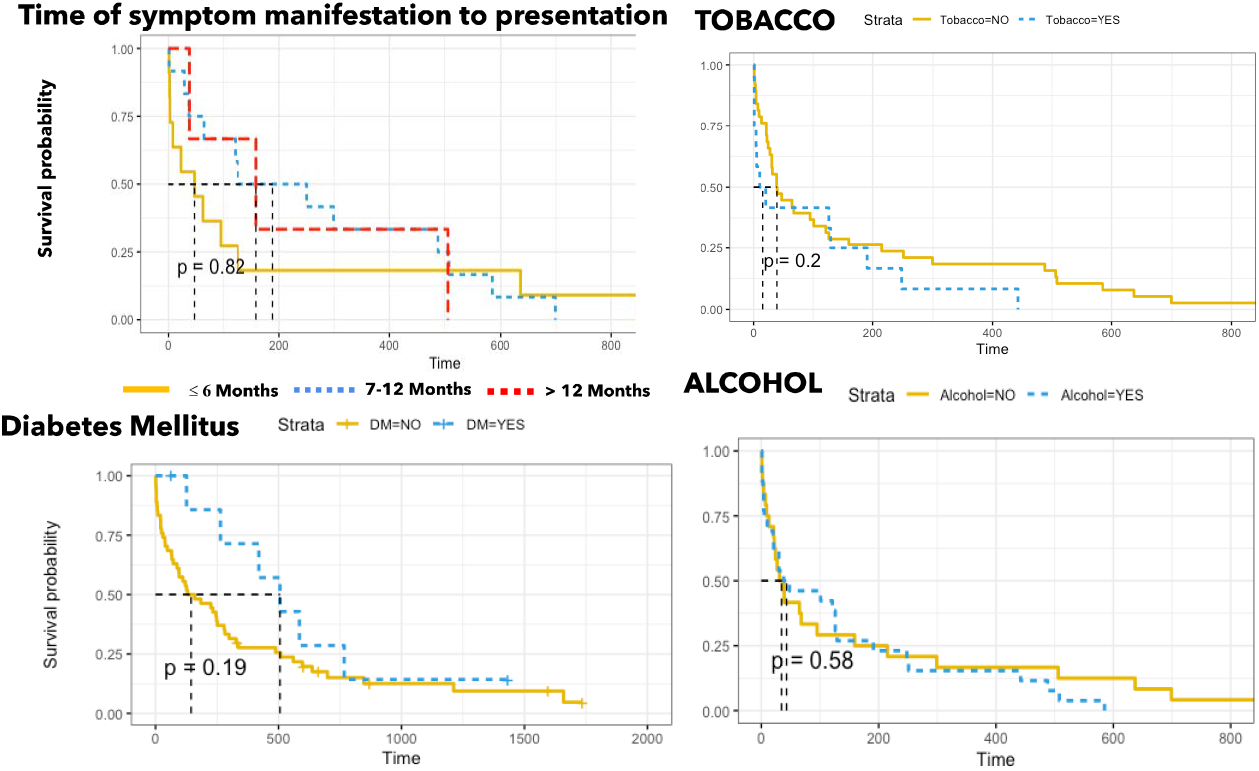
Survival analysis of CRC patients based on time of symptom manifestation to presentation, history of tobacco and alcohol consumption, and diabetes mellitus (Kaplan–Meier curve) Figure 3b shows that patients who presented at the clinic between 7 to 12 months of symptom development had higher mean survival rate (207.0 ± 55.3/192.2 days) compared with those presented at > 12 (197.5 ± 34.9/172.6 days) and ≤ 6 months (182.6 ± 35.1/45.3 days) at p= 0.820. It also patients without a history of tobacco use and alcohol consumption lived longer (mean/median= 162.8 ± 37.2/39 days and 159.5 ± 50.0/35 days) than those with a history of the lifestyles (98.2 ± 38.3/15 days and 135.8 ± 34.3/43 days) at p= 0.200 and 0.580, respectively. Surprisingly, the mean/median survival rate among patients with DM (n= 12) were higher (629.0 ± 185.7/506.0 days) compared with those without the disease (373.2 ± 71.2/145 days, respectively) at p= 0.190.

### Pre- and Post-treatment biomarker values

Overall, significant direct relationships were observed between pre-T and post-T in terms of % lymphocyte (33.05 ± 5.02 and 28.47 ± 4.18), NLR (2.33 ± 0.46 and 3.17 ± 0.69), and PNLR (967.00 ± 302.98 and 1307 ± 360.61) values at p= 0.01, 0.02, and 0.015, respectively. Insignificant direct relationships were observed between pre-T and post-T values of TWBC counts (9.44 ± 0.97 and 12.57 ± 2.71), Neutrophil (51.99 ± 5.38 and 48.93 ± 6.92), platelet (375.42 ± 49.07 and 360.21 ± 32.02), NLPR (0.93 ± 0.20 and 1.09 ± 0.28), and LMR (10.10 ± 1.89 and 5.00 ± 1.33) at p= 0.958, 0.135, 0.085, 0.121, and 0.813, respectively. An indirect negative correlation was observed between pre-T and post-T PLR (195.4 ± 27.75 and 216.7 ± 40.85) at p= 0.844. The pre-T/post-T median value of TWBC, NLR, PLR, PNLR, NLPR, and LMR were 7.00/6.77, 1.50/1.99, 148.3/149.5, 625.6/860.1, 0.59/0.69, and 6.10/3.27, respectively. The post-T LMR and NLPR significantly decreased and increased among patients without metastatic tumours compared with their pre-T values, respectively at p< 0.05 (table 4).

**Table 3:**
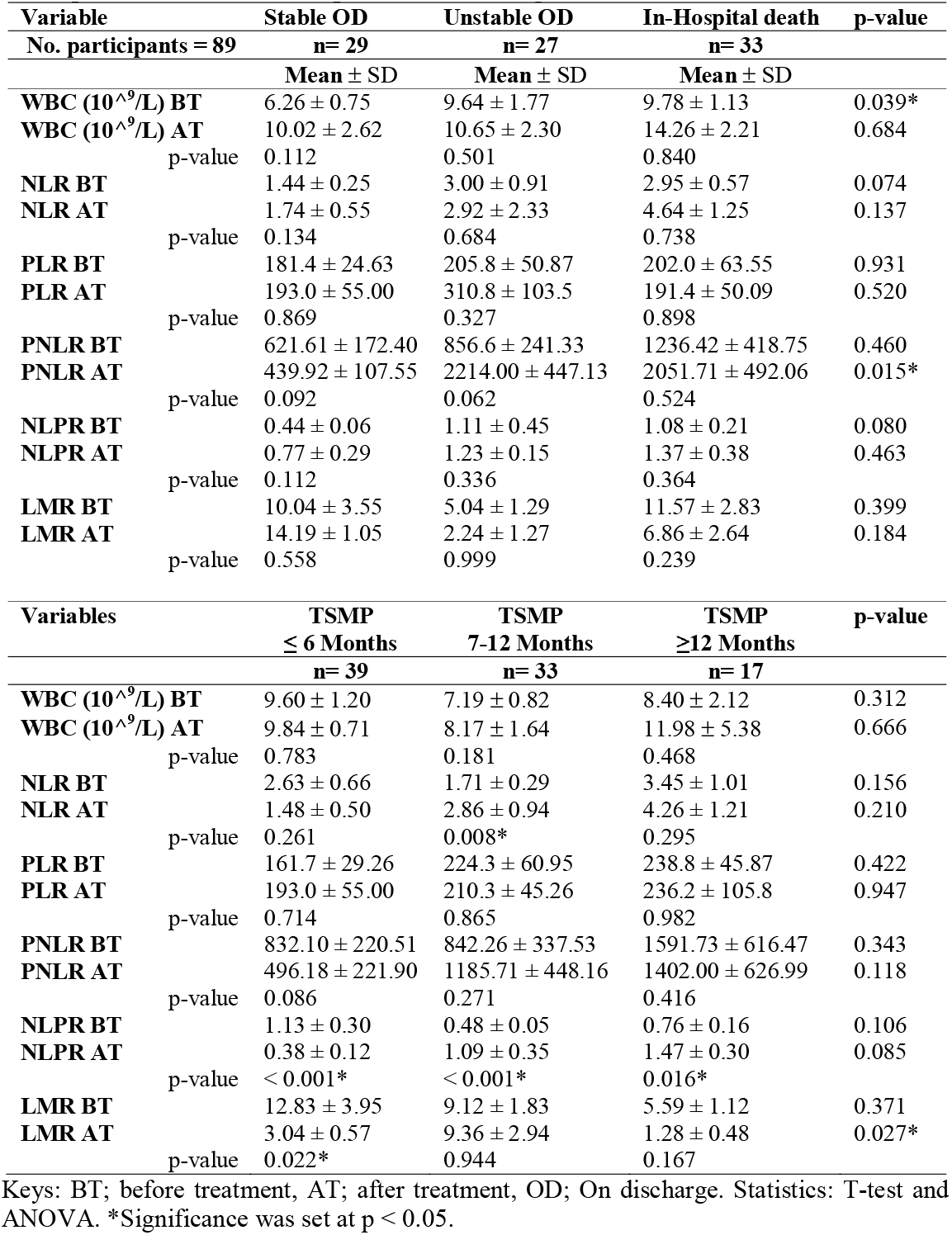
Comparative analysis of hematological indices based on time of symptom development and condition of patient on discharge.

**Table 4:**
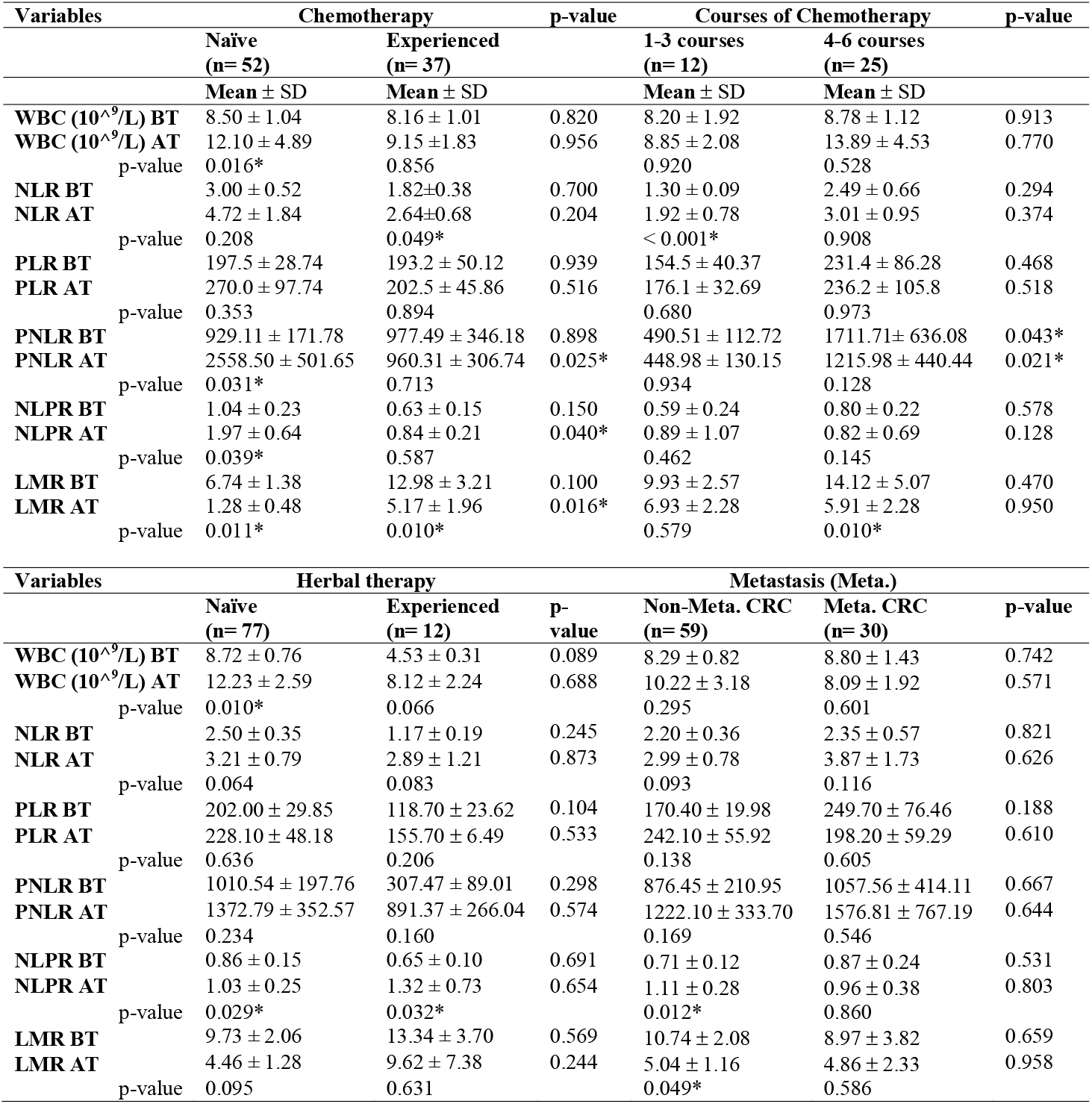
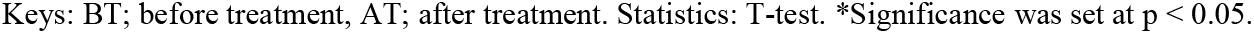
Comparative analysis of hematological indices based on chemo/herbal therapy experience and metastasis.

Irrespective of chemotherapy status, cases of in-hospital death had higher pre-T TWBC counts/ neutrophils compared with patients who were unstable and stable on discharge (9.78 ± 1.13/60.12 ± 4.84, 9.64 ± 1.77/58.56 ± 6.98, and 6.26 ± 0.75/39.47 ± 4.92 at p= 0.039/0.012, respectively). The post-T PNLR of patients who were stable on discharge was approximately 4 times lower than the values of those who died in the hospital or were unstable on discharge at p< 0.05 (table 3). The survivors had a lower pre-T PNLR and NLPR (233.95 ± 118.42 and 0.31 ± 0.05) compared with patients who died out of hospital (943.24 ± 311.83 and 0.99 ± 0.28) and in hospital (1236.42 ± 418.75 and 1.37 ± 0.38) at p= 0.588 and 0.633, respectively. The patient who presented at the clinic > 12 months after symptom manifestation had lower LMR compared with other groups (p< 0.05).

The post-T NLPR of patients who presented ≤ 6 months of symptom manifestation was approximately 3 times lower than their pre-T value (p< 0.05) whereas the post-T PNLR of patients who presented at the clinic > 6 months were higher than their pre-T PNLR at p< 0.05 (table 3). There was a significant direct relationship between age and NLPR (p= 0.019), mortality, and NLPR (p= 0.031).

### Chemotherapy and herbal therapy assessment

Chemotherapy uptake was 41.6%; metastatic CRC (42.3%) and non-metastatic CRC (41.2%). There was a direct relationship between the pre-and post-chemotherapy PNLR (p= 0.000), especially among metastatic cases (p= 0.012). Among chemo-experienced patients, pre-T/post-T median values of TWBC (6.6/6.1), NLR (1.18/2.11), PLR (146.16/153.0), PNLR (357.9/1065), NLPR (0.42/0.69), and LMR (7.84/3.70) were identified.

Among chemo-naïve patients, the pre-T/post-T median values of TWBC (7.7/6.77), NLR (2.02/1.99), PLR (158.61/194.5), PNLR (711.1/693.1), NLPR (0.74/0.65), and LMR (4.69/1.43) were also assessed. Among chemo-naïve patients, the pre-T to post-T TWBC, PNLR, and NLPR significantly increased while the post-T LMR reduced at p< 0.05 (Table 4). Among chemo-experienced patients, the post-T NLR and LMR significantly increased and decreased, respectively compared with the pre-T values (p< 0.05). The post-T PNLR and NLPR were approximately 2.7 and 2.3 times higher while the post-T LMR was 4 times lower among chemo-naïve patients compared with the post-T values among chemo-experienced patients (p< 0.05). The pre-T and post-T PNLR of patients who received 4 to 6 courses of chemotherapy were 3.5 and 2.7 times higher than those who received ≤ 3 courses of chemotherapy (p< 0.05). The post-T LMR of patients who received 4 to 6 courses of chemotherapy was 2.4 times lower compared with their pre-T LMR (p< 0.05). The post-T NLR of patients who received ≤ 3 courses of chemotherapy were significantly reduced compared with their pre-T NLR (p< 0.05). The post-T total WBC and NLPR were significantly increased among patients without a history of herbal therapy compared with their pre-T values (p< 0.05). Only the post-T NLPR was significantly increased among patients with a history of herbal therapy compared with their pre-T values (p< 0.05).

### Chemo-resistance and chemo-sensitive cases

Only 54.1% (n= 20) of the chemotherapy-experienced patients had 6 courses of chemotherapy; chemo-resistant patients (Chemo-R.; 30%) = 6 and chemo-sensitive (Chemo-S.; 70%)= 14. The post-T NLR, PLR, PNLR, and NLPR of patients with chemo-R. tumours were 2.4, 2.3, 1.5, and 3.3 times higher than the post-T values among patients with chemo-S. tumours at p= 0.027, 0.015, 0.022, and 0.131, respectively. The pre-T NLPR of patients with chemo-R. tumours were approximately 2 times higher than that of patients with chemo-S. tumours (p= 0.004). The pre-T and post-T LMR of patients with chemo-R. tumours were 1.9 and 1.6 times lower than that of patients with chemo-S. tumours at p= 0.078 and 0.456, respectively (figure 4).

**Figure 4:**
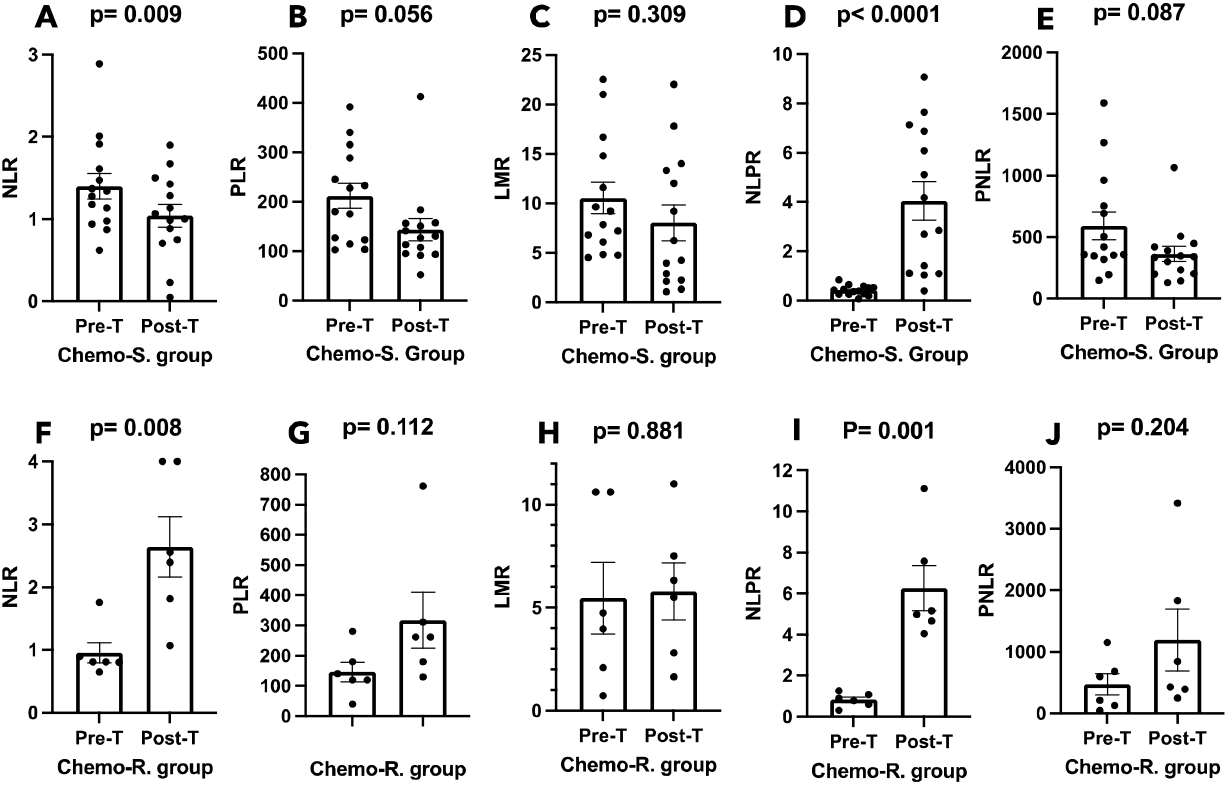
Hematological indices among patients with chemo-sensitive and chemo-Resistant tumours. Among chemo-sensitive (Chemo-S.) patients, figures 4A and 4D show significantly higher pre-treatment (Pre-T) NLR and post-treatment (Post-T) NLPR compared with post-treatment NLR and pre-treatment NLPR (p< 0.05). Among chemo-resistant (Chemo-R.) patients, figures 4F and 4I show significantly higher post-treatment NLR and NLPR values compared with pre-treatment NLR and NLPR (p< 0.05).

## Discussion

In this study, we analyzed the levels of systemic immune-inflammatory indices as an alternative and cost-effective tool for monitoring CRC patients at high mortality risk, especially those in low/medium HDI countries. First, this study revealed that the frequency of the diagnosis and mortality rate significantly increased among the former at the peak of the COVID-19 pandemic, 2020, possibly due to limited access to health facilities at the time. This study also revealed that the disease was more prevalent among patients above the age of 50 years (58%) than in patients who were 50 years and below. This aligns with the study carried out by Alatise et al. and by Saluja et al. who investigated 347 and 160 cases of CRC in Western Nigeria and observed that the disease was dominant among patients aged > 50 years (62.8% and >50%, respectively).^3,14^ The findings of this study is at variance with two other studies carried out in Northern Nigeria, one of 50 cases and one of 605 cases, reported 72% and 62.6% disease dominance among patients aged ≤ 50 years.^15,16^

Regarding tumours site, this study revealed a lower frequency of RCCs (48.1%) compared to LCCs (51.9%). This is discordant with the findings of Edino et al. and Theyra-Enias et al. who reported a high frequency of RCC (77.8% and 55.6%) compared with LCC (22.2% and 44.4%) in Northern Nigeria from 1999 to 2015.^15,17^ This study is also at variance with the findings of Alatise et al. who observed a high frequency of RCC (80%) in Western Nigeria compared with LCC (20%).^3^ Whereas the findings of this study in terms of tumours sites align with the findings of Saluja et al. who reported a lower frequency of RCC (47.6%) compared with LCC (52.4%) in Western Nigeria.^14^ This suggests that the CRCs in Northern Nigeria are more lethal than those observed in Southern or Western Nigeria. The reason for the variation and similarities between the regions is unknown but it could be related to diet, lifestyle, or rate of genetic mutations.^18^

The findings of Irabor et al. (2017) are very similar to the findings of this study in terms of sex and age-related differences in tumour site and grade. They reported a lower RCC frequency but higher frequencies of poorly differentiated adenocarcinoma and rectal tumours among patients aged ≤ 50 years (15%, 43%, and 72%) compared with their > 50 years counterparts (32%, 27%, and 59%, respectively). Additionally, Irabor et al. reported a higher frequency of the disease among females and males who were aged ≤ 50 years (62%) and > 50 years (56%), respectively.^6^ The prevalent tumours grade in the study carried out by Edino et al. and Alatise et al. were poorly differentiated adenocarcinoma (34%) and moderately differentiated adenocarcinoma (55.3%), respectively whereas well-differentiated adenocarcinoma was prevalent among the patients of this study.^3,15^ The high frequency of rectal tumours and poorly differentiated adenocarcinoma among patients aged ≤ 50 years could be due to a high frequency of history of tobacco use, and alcohol consumption. This may affect the clinical outcomes of the patients. Previous studies show that RCC is associated with lower survival compared with LCC due to its metastatic potential, microbiome changes, and high microsatellite instability; MSI-high.^6,19^ This might be the reason for the higher rate of metastasis among patients older than 50 years in this study.

The number of patients who received chemotherapy in this study is lower than the frequency recorded by Edino et al. and Theyra-Enias et al. in Northern Nigeria (94% and 73%, respectively),^15,17^ and the frequency of 67.5% and 50.5% recorded by Saluja et al. and Sharma et al. in Western Nigeria.^14,20^ The reasons for low chemotherapy uptake are due to patients’ reasons, side effects, and lack of funds.^17^ Even though our findings and that of Edino et al. are fourteen years apart, both studies revealed that most of the CRC patients in Nigeria present after 6 months of symptom development.^15^ This might be the explanation for the higher frequency of metastasis among our cohort and high mortality among Nigerian patients. This suggests that the level of awareness and knowledge of the disease is quite low.

Based on the time of presentation at the clinic, the NLR value shows that patients who presented after 6 months of symptom manifestation were at a higher risk of mortality at presentation than patients who presented within 6 months of symptom manifestation. Even though most of the patients aged > 50 years presented at the clinic within 6 months of symptom development, they had a higher frequency of metastasis and stage III/IV CRC compared with their ≤ 50 years counterparts. Despite surgical resections in this group, the three-fold reduction in post-T NLPR was mitigated by the four-fold decrease in post-T LMR. The significant increase in post-T total WBC, neutrophil, monocytes, NLPR, and a significant decrease in lymphocyte, and LMR suggests that patients aged > 50 years responded poorly to treatment due to age-related physiologic limitations. In China, patients with NLR□>□2.72, PLR□>□219.00, and LMR□≤□2.83 were significantly associated with decreased OS and DFS.^21^ In this study, these features were seen among patients aged > 50 years. Thus, the low uptake of chemotherapy by patients aged > 50 years may be responsible for the high mortality rate in 2020 among patients older than 50 years. More so, the high NLR and low LMR among our cohort could be the reason for the high in-hospital death in this study.

In this study, the change from pre-chemotherapy to post-chemotherapy NLR among neoadjuvant chemotherapy-experienced patients (31.1%) is higher than the value reported by Lai et al.^22^ Of note, patients who received more than four courses of chemotherapy had higher pre-T (3.5 times) and post-T (2.7 times) PNLR compared with patients who received three or fewer chemotherapy courses. This suggests that some patients in this study responded poorly to the chemotherapy regimen. This suggests that high PNLR could be used to predict chemotherapy response. Based on symptom alleviation, patients who received six cycles of chemotherapy were classified into subgroups: chemosensitive and chemoresistance. Lower post-chemotherapy NLR, PLR, PNLR, and LMR compared with pre-chemotherapy values were observed among chemosensitive patients whereas the reverse was the case among chemoresistant patients, except for LMR. The pre-and post-chemotherapy LMR were approximately 1.9 and 1.6 times lower among chemoresistant patients compared with chemosensitive patients. Studies have shown that high or persistent elevated preoperative and postoperative NLRs, especially during adjuvant chemotherapy, are strong independent indicators of poor prognosis in patients with stage II or III gastric cancer and colorectal liver metastases.^23-25^ Lai et al. reported that a > 21.5% change in NLR from pre□to post-neoadjuvant chemoradiotherapy was associated with a poor response.^22^ Li et al. reported that LMR is an independent prognostic factor of OS while NLR and LMR were independent prognostic factors of DFS.^21^

Patients with a history of herbal therapy had higher survival rates, lower pre-T TWBC, NLR, PLR, PNLR, NLPR, and higher LMR including lower post-T TWBC, NLR, PLR, PNLR, and higher LMR. Since the mean and median ages of patients who consumed herbal therapy for their ailment before presentation at the clinic were 55.9 years and 54 years, it could be argued that herbal therapy improved patients’ response to chemotherapy through immuno-inflammatory modulation. Further studies are needed to determine the factors responsible for the chemoresistance. A limitation exists here because we also could not analyze and identify the individual constituents of the herbal therapy consumed by the patients before presentation at the clinic. The other limitation of this study is the small-sized subgroups, especially the herbal therapy-experienced subgroup, chemoresistant and chemosensitive subgroups, and the chemotherapy-experienced subgroups (1-3 verses 4-6 courses).

## Conclusion

This study unveils compelling findings regarding patients diagnosed with CRC in Southern Nigeria. It shows that age is a strong driver of disease aggressiveness among patients in Southern Nigeria. It revealed that NLR, PLR, LMR, and PNLR values provide affordable insights into treatment outcomes, especially among elderly patients with late-stage diseases. Interestingly, it shows that herbal therapy uptake prior to presentation influences clinical outcomes and should be investigated further and standardized for possible integration into patient care. There is a need for increased awareness and closer follow-up among patients aged 50 years and above. Due to early-on-set CRC in West Africa, screening for CRC should begin at age ≥ 30 years.

## Data Availability

All data produced in the present work are contained in the manuscript

